# How to improve polygenic prediction from whole-genome sequencing data by leveraging predicted epigenomic features?

**DOI:** 10.1101/2024.10.04.24314860

**Authors:** Wanwen Zeng, Hanmin Guo, Qiao Liu, Wing Hung Wong

## Abstract

Polygenic risk scores (PRS) are crucial in genetics for predicting individual susceptibility to complex diseases by aggregating the effects of numerous genetic variants. Whole-genome sequencing (WGS) has revolutionized our ability to detect rare and even *de novo* variants, creating an exciting opportunity for developing new PRS methods that can effectively leverage rare variants and capture the complex relationships among different variants. Furthermore, regulatory mechanisms play a crucial role in gene expression and disease manifestation, offering avenues to further enhance the performance and interpretation of PRS predictions. Through simulation studies, we highlighted aspects where current PRS methods face challenges when applied to WGS data, aiming to shed light on potential opportunities for further improvement. To address these challenges, we developed Epi-PRS, an approach that leverages the power of genomic large language models (LLM) to impute epigenomic signals across diverse cellular contexts, for use as intermediate variables between genotype and phenotype. A pretrained LLM is employed to transform genotypes into epigenomic signals using personal diploid sequences as inputs, and the genetic risk is then estimated based on the imputed personal epigenomic signals. Epi-PRS enhances the assessment of personal variant impacts, enabling a comprehensive and holistic consideration of genotypic and regulatory information within large genomic regions. Our simulation results demonstrated that incorporating the nuanced effects of non-linear models, rare variants, and regulatory information can provide more precise PRS prediction and better understanding of genetic risk. Applying Epi-PRS to real data from the UK Biobank, our results further showed that Epi-PRS significantly outperforms existing PRS methods in two major diseases: breast cancer and diabetes. This study suggests that PRS methods can benefit from incorporating non-linear models, rare variants, and regulatory information, highlighting the potential for significant advancements in disease risk modeling and enhancing the understanding of precision medicine.

**Significance Statement:** Epi-PRS improves polygenic risk scoring by integrating genomic large language models (LLMs) to impute epigenomic signals as intermediaries between genotype and phenotype. This approach enables a more comprehensive assessment of personal variant impacts by incorporating non-linear models, rare variants, and regulatory mechanisms. By leveraging the power of genomic LLM trained on massive amount of reference epigenomics data, Epi-PRS has demonstrated superior performance over existing PRS methods in predicting genetic risk for breast cancer and diabetes in UK Biobank data. These results highlight the potential of Epi-PRS to improve disease risk modeling and advance the field of precision medicine.

## Introduction

Polygenic risk score (PRS) has become pivotal in genetics for assessing an individual’s susceptibility to complex diseases (1-4). In personalized medicine, PRS enables stratification of individuals based on their genetic risk, facilitating targeted interventions and optimizing clinical outcomes (5, 6). This predictive capability is crucial for early disease detection and prevention, potentially reducing healthcare costs and improving personalized health care (7, 8). Furthermore, PRS elucidates the genetic architecture of complex traits, providing insights into the relevant biological pathways involved in disease etiology (9, 10), which may potentially guide the development of new therapeutic strategies and enhance our ability to identify potential drug targets (11, 12).

Detailed knowledge of how diversity in the human genome sequence affects phenotypic diversity relies on a comprehensive characterization of both sequences and phenotypic variations (13). Whole genome sequencing (WGS) has advanced this field by enabling the detection of numerous rare and even *de novo* non-coding variants (14). WGS is more comprehensive than genotyping arrays and whole exome sequencing (WES) as it allows for the detection of a broader range of variant types and provides more uniform coverage of the entire genome, including regulatory regions (15). Recently, very large scale WGS data with associated clinical phenotypes are being made available for research. These include WGS data for about 500,000 participants from the UK Biobank^16^ and 150,000 participants from the US Million Veterans Program (16, 17). Clearly, there is an urgent need for PRS methods capable of utilizing the full information in WGS data.

Current methods for PRS prediction, such as LDpred2 (18, 19) and PRS-CS (20), have been instrumental in advancing our understanding of genetic risk and have provided invaluable insights into disease susceptibility. These methods were designed with the best available data at the time of their development, typically from SNP arrays that primarily capture common variants (1). As such, they have been highly effective within those contexts. However, with the advent of WGS and larger datasets, the landscape has evolved, revealing new opportunities for refinement. Firstly, many PRS methods rely on linear models, which were well-suited to the data of earlier studies, providing computational efficiency and ease of interpretation. However, linear models assume additive effects of genetic variants and may not fully capture the complex interactions between variants, even as the larger datasets offer rich opportunities to explore these interactions (21, 22). Thus, there is a need to develop non-linear models to account for non-additive effects in larger datasets (23). Secondly, given their focus on common variants, traditional PRS methods were not designed to incorporate rare and *de novo* variants. These rare variants, though less frequent, can have substantial impacts on disease risk (24), and incorporating them could enhance the predictive power of PRS (25, 26). Finally, the role of regulatory mechanisms in gene expression and disease manifestation has become increasingly clear (27-30). For example, genetic variants can influence disease risk not only through their direct effects on protein function but also by altering the activities of regulatory elements such as enhancers, promoters, and transcription factor binding sites (31-33). Current PRS methods either ignore this regulatory information (19, 20) or incorporate as linear model priors and parameters (34, 35). As our understanding of regulatory mechanisms expands, there are exciting opportunities to improve PRS models by integrating this critical layer of biological information (34, 36, 37).

To investigate the factors limiting the accuracy of current PRS methods, we conducted comprehensive simulation studies. First, we examined the limits imposed by linear models. Our simulations revealed that when diseases are influenced by complex interactions between genetic variants, non-linear models can significantly outperform traditional linear models by capturing these intricate interactions and improving predictive performance. Second, we explored the role of rare variants. The simulation showed that current PRS methods are not effective in using the information from rare variants. Therefore, they performed poorly in predicting genetic risk for diseases where rare variants play a critical role.

To address these challenges, we developed Epi-PRS, a non-linear method for polygenic prediction capable of using information from both common and rare variants. Our approach is to use the phased personal sequences to represent all variants in large genome regions, regardless of whether the variants are common, rare or *de novo*. Specifically, for every individual in our study cohort, we use a genomic large language model (LLM) to transform the phased DNA sequences into a set of context-specific and region-specific epigenomic features. Then, we use these personal epigenomic features as explanatory variables to learn a nonlinear polygenic risk prediction model. This approach is motivated by the important role of regulatory mechanisms in disease etiology (27-30). For example, genetic variants can influence disease risk by altering the activities of regulatory elements such as enhancers, promoters, and transcription factor binding sites (31-33). With the advent of genomic LLMs that are learned from a massive amount of epigenomic data sets across diverse cellular contexts, we can now predict many types of epigenomic signals across a genomic region based on the DNA sequence of that region. By leveraging these LLMs, our approach offers a principled way to incorporate the effects of all variants via their impact on predicted epigenomic features. Moreover, the epigenomic features are biologically more meaningful as explanatory variables than the genotypes themselves. Thus disease risk models based on predicted epigenomic features may offer deeper insight on the molecular basis of diseases. Our simulation studies demonstrated that this approach (Epi-PRS) significantly outperforms traditional PRS methods. Importantly, when applied to UK Biobank data on breast cancer and type 2 diabetes, Epi-PRS showed clear and substantial improvements over state-of-the-art PRS methods, highlighting its potential for enhancing disease risk prediction through the integration of advanced computational techniques and comprehensive genetic insights.

In summary, this study not only introduces a method for enhancing polygenic risk prediction but also provides new insights into the factors affecting the performance of PRS methods. The simulation results suggest that existing PRS methods can be further strengthened by integrating non-linear models, rare variants, and regulatory information, paving the way to significant advancements in disease risk modeling. Our findings provide useful perspectives on how to use WGS data to overcome the limitations of current PRS methods. It is hoped that our results can enable more precise genetic risk assessments and deeper understanding of the regulatory mechanisms underlying complex diseases.

## Results

### Overview of the Epi-PRS model

The Epi-PRS workflow introduces a comprehensive approach to disease risk prediction by integrating personal genomic and epigenomic data in novel ways. As shown in Figure 1, Epi-PRS models how personal genotypes influence phenotypes through a diverse array of genomic and epigenomic profiles, imputed by a genomic large language model (LLM) trained on reference data from a diverse range of cellular contexts. A distinguishing feature of Epi-PRS is its ability to handle diploid sequences, allowing for a more comprehensive assessment of variant interactions in personal DNA sequences, which enhances the accuracy of disease risk predictions. The Epi-PRS workflow comprises three major steps: personal genome construction, epigenomic feature extraction, and disease risk prediction. Detailed descriptions of the methods are provided in the Methods section.

**Figure 1.**
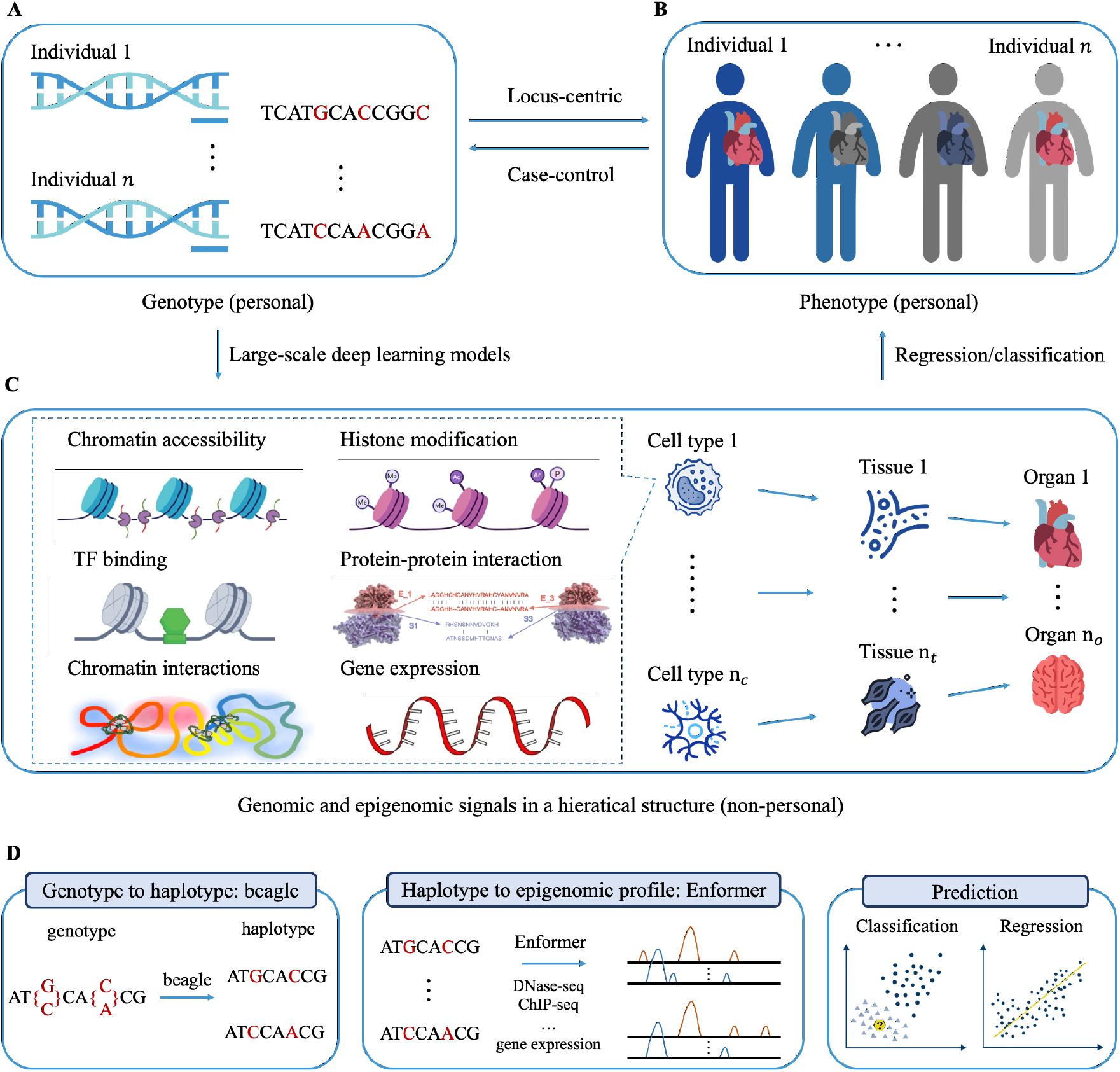
A simplified diagram that shows the understanding of how A) personal genotypes affect B) personal phenotypes, which requires C) the modeling of the relationship between different layers of omics based on non-personal context-specific reference data using large-scale deep learning models. D) Assuming WGS and phenotype data are available from the cohort, the main steps of Epi-PRS. 1) Personal genome construction: maternal and paternal genome will be constructed from personal WGS as the input for Epi-PRS. 2) Epigenomic features extraction: genomic LLM (e.g., Enformer) will be applied to haploid genome to obtain the personal epigenomic features. 3) Risk prediction: build model for disease risk as a function of the epigenomic features.

Epi-PRS offers several unique aspects that extend beyond traditional PRS prediction approaches: 1) Integration of molecular phenotypes: While conventional methods focus primarily on genotype data, Epi-PRS incorporates imputed molecular phenotypes, providing additional context for how genetic variants may influence disease risk through regulatory mechanisms. 2) Diploid sequence modeling: Epi-PRS captures the combinatorial effects of multiple variants by processing diploid sequences, which is important for understanding the complexity of polygenic traits. 3) Inclusion of rare and de novo variants: Epi-PRS is able to include rare and *de novo* variants, expanding the scope of genetic information considered in disease risk prediction. These advancements build on the strong foundation laid by existing PRS methods and aim to provide additional perspectives for improving disease risk prediction.

### PRS prediction on simulation data

A series of simulation experiments were carefully designed and conducted to evaluate the performance of PRS methods. The evaluation focused on three key comparisons: 1) the efficacy of linear models versus non-linear models; 2) incorporating regulatory information or not; and 3) considering the impact of rare variants or not. These comprehensive comparisons aimed to provide a deeper understanding of the factors influencing PRS accuracy and effectiveness. A regulatory-based phenotype simulation tool was developed to ensure that the simulated phenotypes are affected by different components, including 1) epigenetic effects, 2) SNP direct effects, 3) environmental effects and 4) SNP-interaction effects as described in the Methods section. To evaluate the power gained from inclusion of rare variants, the simulation tool can select the proportion of variants to be rare. Phenotypes were simulated based on different linkage disequilibrium (LD) blocks, incorporating both common and rare variants. Various genetic architectures were considered, with different proportions of rare and common variants and different proportions of epigenetic and SNP direct effects to simulate scenarios where regulatory elements influence gene expression and subsequently disease risk. Following other PRS methods (20), simulation studies used real genetic data from the UK Biobank European ancestry samples (N = 20,000) in 2 LD blocks: chr6:192074807-21684054 and chr6:31571218-32682664. Phenotypes were generated as a linear combination of the above four types of effects with added noise as explained in Methods section.

The simulations were used to evaluate Epi-PRS as well as several traditional PRS methods, including: 1) LDpred2: A widely used Bayesian approach for PRS estimation. 2) PRS-CS: Another linear-model based method that utilizes continuous shrinkage priors to account for LD and effect size heterogeneity. 3) Genotype - GBRT: To evaluate if non-linear models can improve prediction, we also implemented a Gradient Boosting Regression Trees (GBRT) method to predict disease risk from all variants in the genotype. 4) Genotype - PCA - GBRT: Performing Principal Component Analysis (PCA) first for all the variants in genotype, then using the PCA output as input for GBRT. The PRS-CS and LDpred2 are linear models that rely on GWAS summary statistics, while Genotype-GBRT and Genotype-PCA-GBRT are non-linear models based on all genetic markers with/without PCA dimension reduction. Since the causal SNPs are located in specific LD blocks, LD block-specific prediction was performed and two LD blocks were selected for testing and comparison of the methods: chr6:192074807-21684054 and chr6:31571218-32682664. The predictive accuracy of the models was assessed using the Area Under the Receiver Operating Characteristic Curve (AUC).

#### 1) The comparison between linear models and non-linear models

To quantify the potential increase in power from the inclusion of non-linear models, a series of simulation experiments varying the number of individuals in the training dataset were conducted. The training dataset sizes were incremented from 1,000 to 2,000, 4,000, 8,000, and 16,000, while the performance was consistently evaluated using a fixed test dataset of 4,000 individuals. In this study, Epi-PRS, Genotype-GBRT, and Genotype-PCA-GBRT were categorized as non-linear models, whereas PRS-CS and LDPred2 were categorized as linear models. In the simulations, the phenotypes will be affected by the SNPs direct effect and SNP-interaction effects while the proportion of epigenetic effects and environmental effects are set to zero. The proportion of rare variants is set to zero, which are the standard simulation setting in previous PRS methods.

Compared with the traditional linear model, we observed a notable gain in power using the non-linear framework, with comparable improvements across larger sample sizes (Figure 2 and Supplementary Table 1, 2). However, when the training dataset is small, non-linear models tend to overfit, resulting in lower performance compared to linear models. This is likely due to the complexity and flexibility of non-linear models, which can capture intricate SNP-interaction effects but may overfit when there are insufficient training instances. As the number of training instances increased, the performance of both linear and non-linear models improved. However, the improvement for non-linear models was more significant compared to linear models. In particular, when the training dataset exceeded 2,000 instances, non-linear models began to outperform linear models significantly. These results indicate that for larger cohorts, non-linear models are more effective and can achieve superior predictive performance. Conversely, in scenarios with limited training data, linear models exhibit better generalization and are less prone to overfitting. Therefore, the choice between linear and non-linear models should be guided by the size of the available training dataset. For extensive datasets, non-linear models are preferable, whereas for smaller datasets, linear models offer better generalizability and reliability.

**Figure 2.**
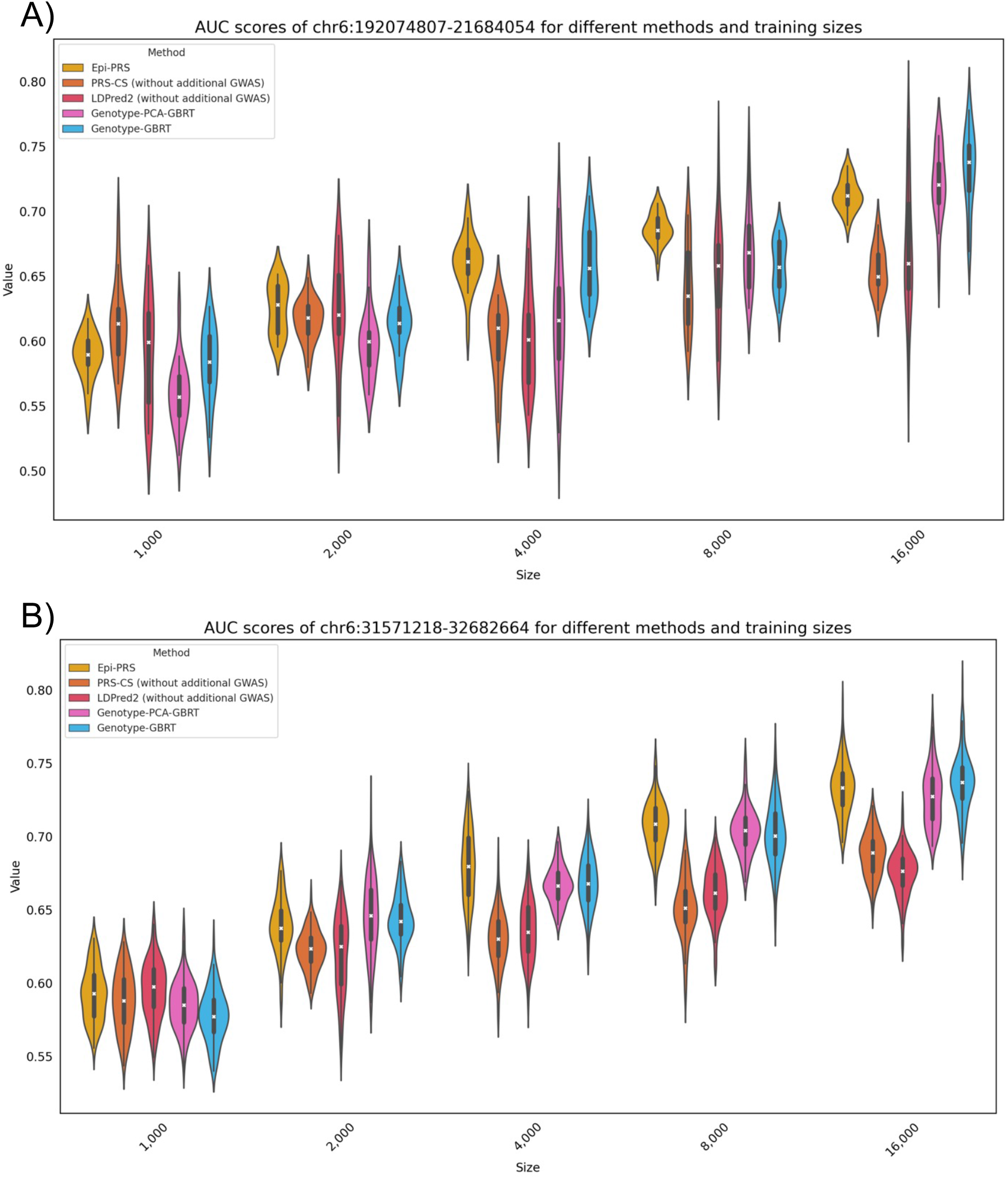
AUC scores for different methods in two LD regions with varying numbers of training data. The violin plot in different color represents the AUC score distribution of a specific method based on the 20 independent simulations. It is seen that the performance of different methods increases when the size of training data also increases. No-linear methods (Epi-PRS, Genotype-PCA-GBRT, and Genotype-GBRT) offers significant advantage over linear methods (LDPred2 and PRS-CS) in large sample size settings.

#### 2) The comparison between whether regulatory information is considered or not

To quantify the potential power gain from considering the effect of regulatory mechanisms, a series of simulation experiments with various proportion of epigenetic effects and fixed proportion of SNP interaction effects were conducted. The proportions of epigenetic effects were varied from 0%, 25%, 50%, 75%, and 100% while the proportion of environmental effects and rare variants were set to 0%. Epi-PRS effectively captured the influence of regulatory mechanisms on gene expression and demonstrated better predictive accuracy by achieving a significantly higher AUC compared to LDpred2 and PRS-CS across all simulation scenarios for the two LD blocks (Figure 3 and Supplementary Table 3, 4). When the epigenetic effect was set to 0% and the SNP direct effect was set to 100%, Epi-PRS, Genotype-GBRT, and Genotype-PCA-GBRT achieve similar AUCs and clearly outperformed traditional PRS methods. On the other hand, there was a clear improvement in the performance of Epi-PRS as the proportion of the epigenetic effect increased, while such improvement was not seen in all the other methods. This suggests that the biggest power gain within the case of the SNPs affecting phenotypes is from epigenetic effect and Epi-PRS can effectively extracting useful regulatory features. When the epigenetic effects were simulated based on regulatory elements in blood cell types, Epi-PRS using blood features (i.e., using only the Enformer predictions for epigenomic data tracks in blood cell types in the training data) further improved the performance compared to Epi-PRS using all features, implying that reducing feature dimension with tissue-specific features is crucial when the disease-related cell types or tissues are known. In contrast, using PCA to reduce the dimension of genotype did not significantly impact the final prediction. These simulation results show that when a disease is affected by epigenetic regulation, Epi-PRS achieves much better performance than existing PRS methods. Furthermore, Epi-PRS power gains are expected to be more dramatic when a better understanding of the disease mechanisms is known and related epigenomic features can be selected.

**Figure 3.**
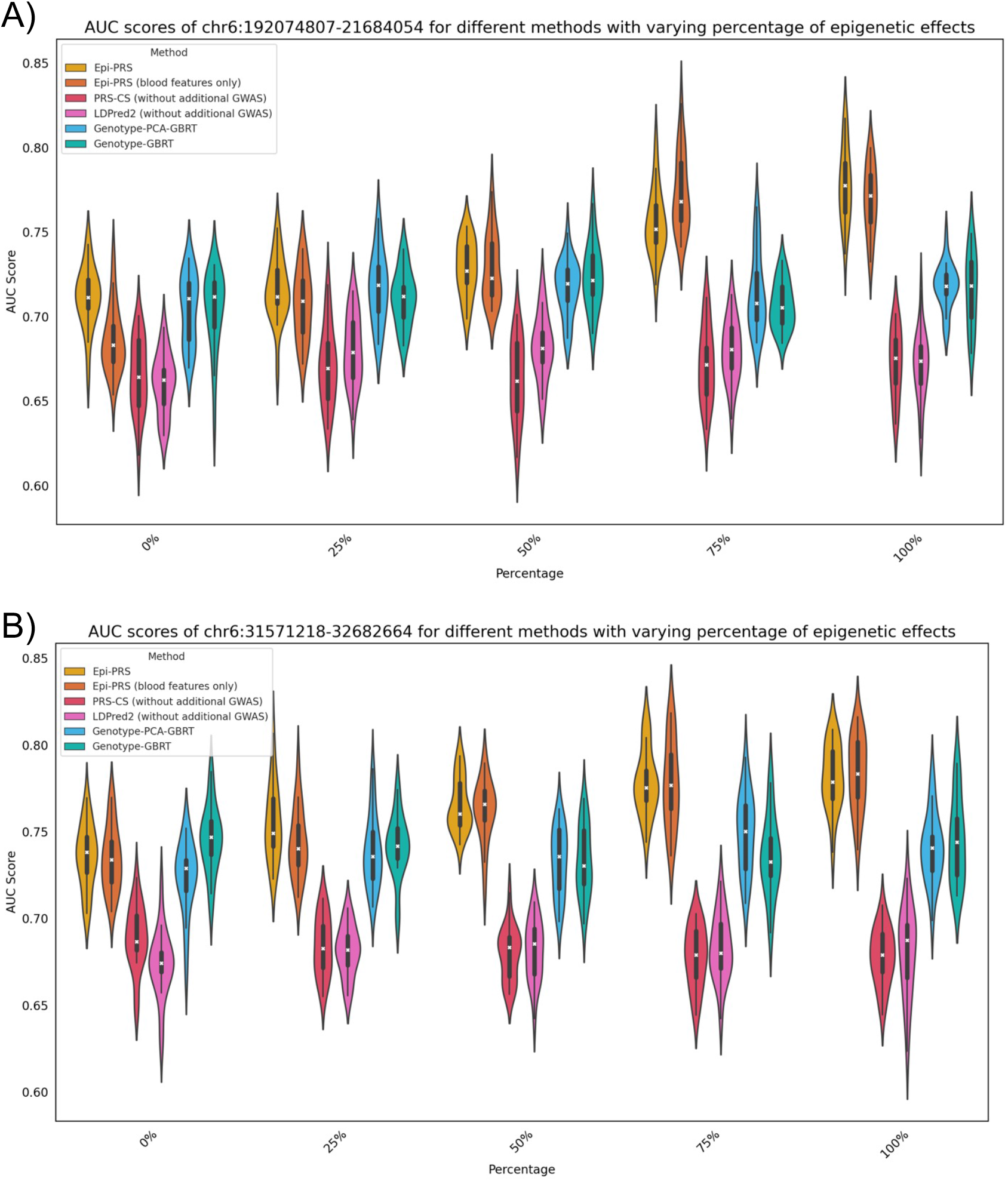
AUC scores for different methods in two LD regions with varying percentages of epigenetic effects. The violin plot in different color represents the AUC score distribution of a specific method based on the 20 independent simulations. Epi-PRS offers significant improvement over other methods especially when the epigenetic effects are large. Reducing dimension of the imputed epigenomic features by only selecting the tissue-specific features leads to notable performance improvement when the disease-related cell types or tissues are known.

#### 3) Comparison results under increasing level of contribution by rare variants

Next, the PRS prediction performance was investigated by varying the proportion of rare variants while setting the proportion of epigenetic effects to zero. Epi-PRS again demonstrated better performance by achieving a 12% higher AUC compared to traditional PRS methods across all simulation scenarios (Figure 4 and Supplementary Table 5, 6). In this setting, the inter-SNP interaction effect was added to simulate the phenotype, a non-linear effect that cannot be captured by linear models. When the proportion of rare variants was set to 0%, Epi-PRS outperformed traditional methods by capturing non-linear effects, leading to an increase in AUC by approximately 11-12%. However, since the simulation was not related to regulatory information, Epi-PRS performed very similarly to Genotype-GBRT and Genotype-PCA-GBRT. As the proportion of rare variants increased, the performance of Epi-PRS improved, while traditional PRS methods (PRS-CS and LDpred2) decreased, and the Genotype-GBRT and Genotype-PCA-GBRT maintained a similar performance level. In scenarios with 75% rare variants, Epi-PRS showed a remarkable improvement in predictive accuracy, with AUC values increased by up to 20% compared to traditional methods, highlighting its ability to leverage rare variant information effectively. As the proportion of rare variants increased to 100%, the traditional methods failed to predict disease risk, while Epi-PRS consistently performed well. In above evaluations, performance was assessed by varying one condition at a time. To further compare the three nonlinear methods, Epi-PRS, Genotype-GBRT, and Genotype-PCA-GBRT were evaluated under combined conditions (50% rare variants and 50% epigenetic effect). The results showed that Epi-PRS achieved an average area under the receiver operating characteristic curve (auROC) of 0.7492, outperforming Genotype-GBRT and Genotype-PCA-GBRT, which had auROCs of 0.6993 and 0.7015, respectively. These comprehensive simulation studies highlight the potential of advanced PRS methods like GBRT and Epi-PRS to significantly advance polygenic risk prediction. In particular, Epi-PRS is anticipated to excel in situations where a significant portion of heritability is primarily driven by rare variants through regulatory effects.

**Figure 4.**
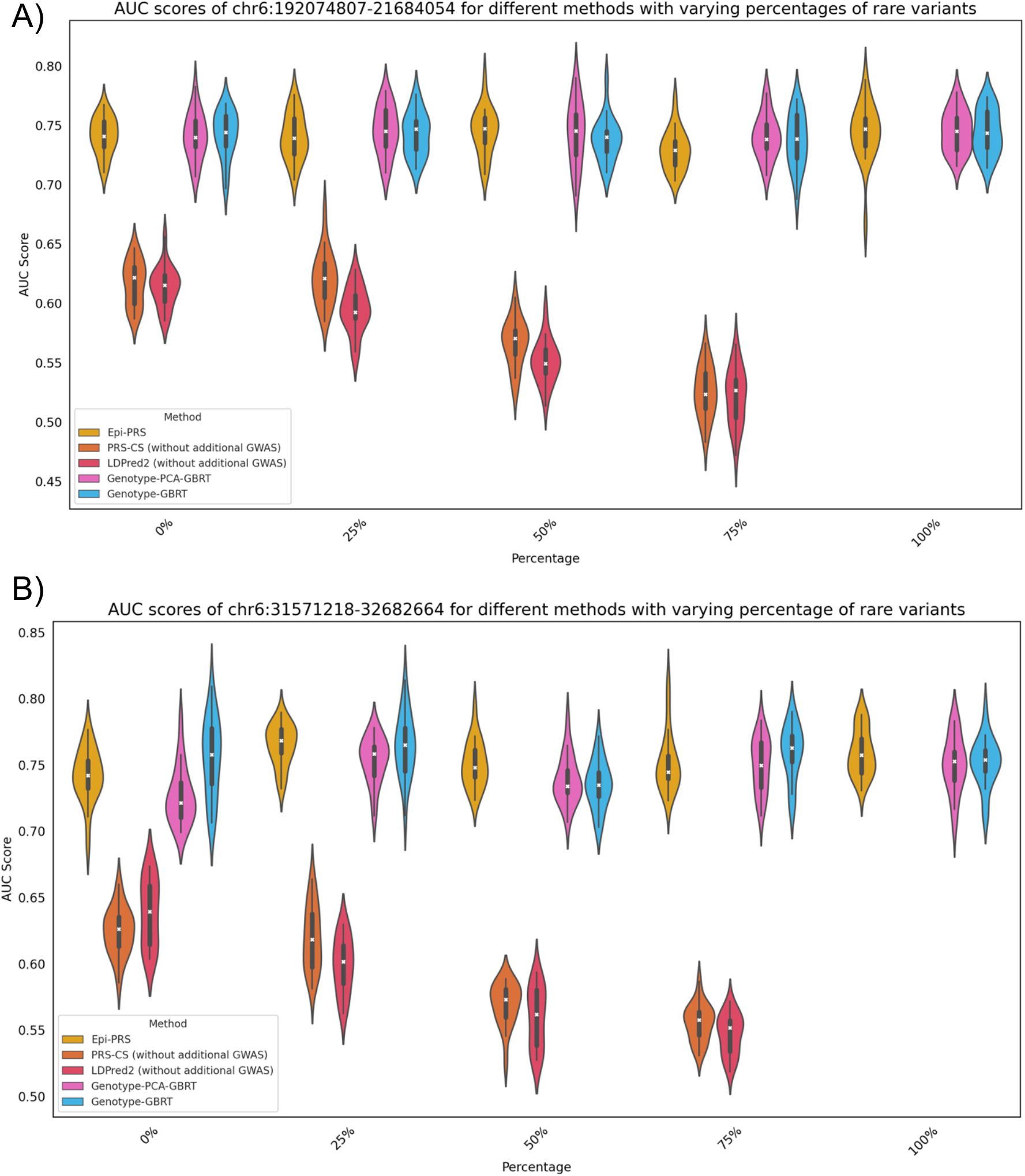
AUC scores for different methods in two LD regions with varying percentages of rare variants. The violin plot in different color represents the AUC score distribution of a specific method based on the 20 independent simulations. Performance of linear methods (LDPred2 and PRS-CS) significantly decrease as the percentage of rare variants increase.

### PRS prediction on the UK Biobank

The Epi-PRS method was applied to the prediction of breast cancer and type 2 diabetes (T2D) using data from the UK Biobank. The performance of a predictor was measured by its relative proportional gain in auROC over random guessing, defined as λ=(A-0.5)/0.5, where A is the auROC of the predictor in the test data. This performance index varies between 0 (for a random guess predictor) and 1 (for a perfect predictor).

#### Breast Cancer

Data were collected from 10,547 breast cancer female subjects (cases) and 10,547 female subjects (controls). Five LD blocks were selected based on a significance threshold applied to variants with a stringent p-value. Since WGS data is not yet available for all subjects, phased SNP genotype data were used to construct the input genomic sequences for epigenomic feature prediction. Initially, the phenotype prediction performance was tested using a single LD block. The dataset was randomly split into an 80% training set and a 20% testing set. The performance of Epi-PRS was compared to three state-of-the-art baseline methods: 1) Genotype-GBRT, 2) LDpred2 (a Bayesian logistic regression method based on SNP genotypes), and 3) PRS-CS (another Bayesian logistic regression method based on SNP genotypes that also utilized external data, such as summary statistics from external GWAS studies using approximately 200,000 samples). As shown in Table 1, Epi-PRS achieved the best performance in each of the five LD blocks, with an auROC ranging from 0.5379 to 0.5917. The improvement was particularly substantial in the fourth LD block. Next, the information from the five LD blocks was combined by concatenating their epigenomic features. The resulting method, “Epi-PRS-concat,” was compared to four baseline methods: 1) LDpred2-WG (applied to the whole genome), 2) PRS-CS-WG (applied to the whole genome), 3) PRS-CS-WG-IN (based on SNPs from the whole genome without using external GWAS summary statistics), and 4) a new combined model. Epi-PRS based on SNPs in the five LD blocks achieved a λ of 0.3994, while the other baseline methods using SNPs from the whole genome achieved λ values between 0.093 and 0.2798 (Figure 5A). The gap between PRS-CS-WG and PRS-CS-WG-IN suggests that the relatively strong performance of PRS-CS-WG is probably due to the extra information provided by external GWAS data.

**Table 1.** The performance of Epi-PRS and baseline methods on polygenic prediction of breast cancer based on single LD block. The AUC of different methods using five different LD blocks on breast cancer. Epi-PRS based on only SNPs in the 5 LD blocks achieves a λ of 0.3994 while the other 4 baseline methods using SNPs in whole genome only achieve λ of 0.093-0.2798. λ = (*A* − 0.5)/0.5, where *A* is the auROC of the predictor in the test data.

| LD block | PRS-CS | LDPred2 | Genotype-GBRT | Epi-PRS |
| --- | --- | --- | --- | --- |
| chr5:55417349-56621102 | 0.5306 | 0.5194 | 0.5239 | **0.5412** |
| chr10:123231465-123900545 | 0.5445 | 0.5479 | 0.5496 | **0.5554** |
| chr11:68005825-69516130 | 0.5157 | 0.5129 | 0.5168 | **0.5392** |
| chr16:52035823-5338257 | 0.5387 | 0.5341 | 0.5316 | **0.5917** |
| chr22:27834752-29651799 | 0.5049 | 0.5082 | 0.5204 | **0.5497** |

**Figure 5.**
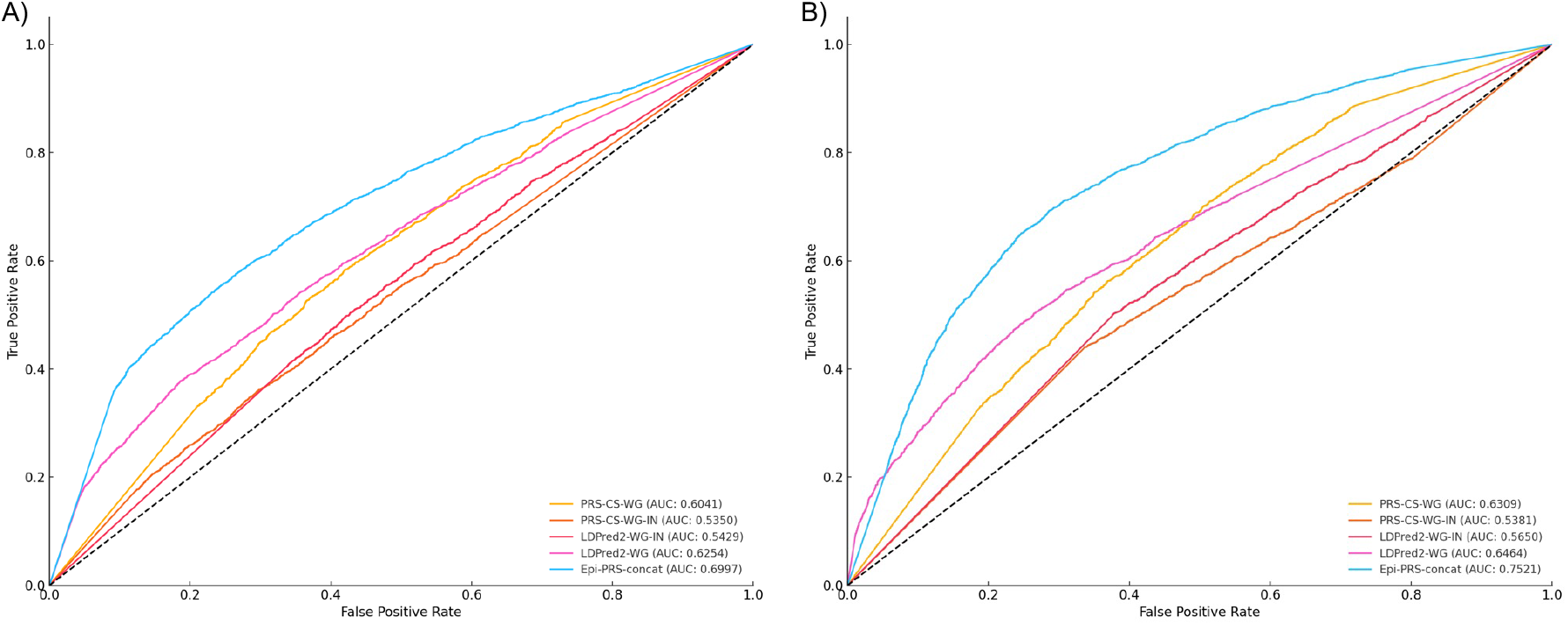
ROC curves of different methods using A) breast cancer and B) type 2 diabetes data from UKBB. Epi-PRS-concat (combining information from multiple LD blocks) outperforms existing methods that utilize whole genome-wide information and external GWAS summary statistics data.

#### T2D

Data were collected from 20,000 case subjects and an equal number of randomly selected control subjects without T2D. Eleven LD blocks were chosen based on a stringent p-value cutoff. Epi-PRS with a single LD block (chr16:53382572-55903774) achieved nearly the best performance compared to four baseline methods that utilized information from all variants across the genome (Table 2). Combining information from multiple LD blocks further improved the prediction performance of Epi-PRS, achieving a λ of 0.515, outperforming the other four baseline methods with λ values ranging from 0.077 to 0.3236 (Figure 5B). This non-linear approach to risk prediction based on epigenomic features significantly improved performance over the baseline methods. These results demonstrate that incorporating haploid sequence context around each variant and considering the effect of personal variants in different cellular contexts is a powerful approach to improving PRS prediction performance. Epi-PRS showed superior performance for both breast cancer and T2D, where regulatory mechanisms play a significant role. The incorporation of epigenomic features from diverse cellular contexts provide a comprehensive assessment of genetic risk, outperforming traditional PRS methods. This highlights the potential of Epi-PRS to advance polygenic prediction and deepen our understanding of the genetic basis of complex diseases.

**Table 2.** The performance of Epi-PRS and baseline methods on polygenic prediction of T2D based on single LD block. The AUC of different methods using eleven different LD blocks. Epi-PRS achieves a λ of 0.515 while other 4 baseline methods only achieve λ of 0.077-0.3236. λ = (*A* − 0.5)/0.5, where *A* is the auROC of the predictor in the test data.

| LD block | PRS-CS | LDPred2 | Genotype-GBRT | Epi-PRS |
| --- | --- | --- | --- | --- |
| chr3:185068255-186890344 | 0.5416 | 0.5235 | 0.5333 | **0.5467** |
| chr4:5502388-6773043 | 0.5387 | 0.5239 | 0.5315 | **0.5548** |
| chr6:19207487-21684065 | 0.5247 | 0.5175 | 0.5386 | **0.5721** |
| chr6:31571218-32682664 | 0.5187 | 0.5129 | 0.5474 | **0.6083** |
| chr6:32682664-33236497 | 0.5257 | 0.5166 | 0.5447 | **0.5574** |
| chr8:116096495-119685457 | 0.5253 | 0.5012 | 0.5287 | **0.6072** |
| chr9:20463534-22206559 | 0.5327 | 0.5281 | 0.5354 | **0.5894** |
| chr10:93335047-95396368 | 0.505 | 0.5052 | 0.5218 | **0.5530** |
| chr10:112561493-115328432 | 0.5604 | 0.5189 | 0.5537 | **0.6229** |
| chr12:3677037-4417679 | 0.5235 | 0.5191 | 0.5343 | **0.5432** |
| chr16:53382572-55903774 | 0.5045 | 0.5067 | 0.5322 | **0.6543** |

## Discussion

With the increasing availability of WGS data, significant challenges and opportunities arise in the field of genetics for improving disease risk prediction. To address these challenges, this study introduces Epi-PRS, an approach that explores several potential improvements to existing PRS methods. These improvements include 1) the adoption of non-linear models: non-linear models have the potential to capture complex interactions between genetic variants that linear models may not detect. By leveraging these non-linear models, the accuracy and predictive power of PRS can potentially be enhanced. Simulation studies in this research demonstrated that non-linear models, such as Epi-PRS, outperform linear models when the training dataset is sufficiently large. This finding suggests that non-linear models are more effective in capturing intricate genetic interactions and can provide superior predictive performance for large cohorts; 2) the consideration of rare variants: rare genetic variants, despite their low frequency, can have substantial impacts on disease risk. Incorporating these variants into PRS calculations can provide a more comprehensive and accurate assessment of genetic risk. Epi-PRS achieves this by doing risk modeling based epigenomic features predicted by a genomic LLM that can integrate information from all the variants, including rare or de novo variants, on the two sequence alleles of a genetic loci. The simulation studies indicated that Epi-PRS may significantly benefit from including rare variants, particularly in scenarios where rare variants play a critical role in disease etiology. In contrast, genotype-based PRS methods, even those using advanced nonlinear predictions, may miss the contributions of rare variants, leading to suboptimal predictions and 3) the integration of regulatory information: regulatory elements play a critical role in gene expression and disease manifestation. By integrating regulatory information into PRS models, deeper insights into the genetic architecture of diseases can be gained, potentially improving the precision of risk predictions. The simulation studies confirmed the importance of considering regulatory mechanisms. This result underscores the value of integrating epigenomic features to enhance the predictive accuracy of PRS models. The comprehensive simulation studies conducted in this research systematically evaluated the impact of non-linear models, rare variants, and regulatory information on PRS prediction. These evaluations provide valuable insights into how each aspect contributes to the performance of PRS methods. By addressing the limitations of existing PRS methodologies, the findings suggest that significant advancements in disease risk modeling can be achieved, ultimately enhancing the understanding of precision medicine. To test Epi-PRS in real data, we apply it to breast cancer and type 2 diabetes data from UK Biobank. The results, based on personal genome sequences imputed from genotyping array data, showed that Epi-PRS outperformed existing PRS methods, confirming the potential for improved disease risk prediction through the integration of non-linear models, rare variants, and regulatory information. This study indicates that incorporating these elements into PRS models can lead to more accurate and comprehensive assessments of genetic risk. Although we were forced to imputed phased genomic sequences because WGS data were not available at the time of this research, the improvement by Epi-PRS over the competing methods should be real. In fact, the improvement would have been even larger if Epi-PRS were based on the real WGS data for the subjects.

While the results of this study are promising, further validation across different diseases is essential to ensure the broad applicability and effectiveness of Epi-PRS. It is possible that the best performance may result from using genotype-only, epigenomic features-only, or a combination of both, depending on the disease of interest. Future research should focus on validating the usefulness of predicted epigenomic features for polygenic prediction in various diseases. Additionally, the integration of other LLMs in place of Enformer could offer further improvements. Future models might provide faster inference, enabling more efficient data processing (38, 39). For example, QUICK can be adopted as they use a group of novel optimized CUDA kernels for the efficient inference of quantized LLMs (40). Another improvement is the high-resolution predictions of epigenomic and genomic profile. By accurately predicting the impact of genetic variants on transcription factor binding at a single base-pair resolution, BPNet enhances the precision of transcription factor binding prediction (41). This model can identify how individual variants alter the binding affinity of transcription factors, offering detailed insights into gene regulation mechanisms. Furthermore, developing models that incorporate cellular context specificity will allow for more accurate modeling of disease-specific regulatory mechanisms. For instance, models like EpiGePT have been designed to predict various types of epigenomic signals given a specific cellular context (42). By integrating such models, we can better understand how genetic variants influence disease risk in a context-dependent manner. This is crucial for diseases that manifest differently across various cell types and tissues, such as cancers and autoimmune disorders.

In summary, this research underscores the importance of integrating advanced computational models, rare variant analysis, and regulatory information to improve PRS methods. These innovations have the potential to elevate the precision of genetic risk predictions and facilitate a deeper understanding of the regulatory mechanisms underlying complex diseases. As a result, the goal of personalized medicine, where treatments and preventive strategies are tailored to the unique genetic makeup of each individual, becomes more attainable.

## Materials and Methods

### Workflow of the Epi-PRS

This workflow is comprised of three major steps: 1) personal genome construction, 2) epigenomic feature extraction, and 3) risk prediction, which are detailed as follows.

#### Personal Genome Construction

The first step in the Epi-PRS workflow involves constructing the maternal and paternal genomes from personal WGS data, which serve as the input for Epi-PRS. This process is detailed in the following steps: A) Variant Filtering: The current version of Epi-PRS focuses exclusively on SNVs. We start by processing the variant call format (VCF) file containing the genetic profiles of all individuals. Using vcftools (43), we filter out all insertions and deletions (INDELs), retaining only the SNPs. B) Genotype Phasing: Next, we phase the genotypes using the reference-free Beagle (44) software. Phasing is essential for distinguishing between the maternal and paternal alleles, which is critical for subsequent steps. C) Personal Genome Construction: Finally, we employ the vcf2diploid (45) tool to reconstruct the maternal and paternal personal genomes for each individual. This step produces two separate haploid genome sequences for each person, representing the maternal and paternal contributions.

#### Epigenomic Feature Extraction

In the second step, we extract epigenomic features using a pretrained genomic LLM, such as Enformer. This process involves several sub-steps: A) Feature Extraction: The genomic LLM (e.g., Enformer) is applied to each haploid genome to predict genomic and epigenomic features, including gene expression, chromatin accessibility, ChIP-seq signals, and histone modification signals across a diverse set of cell lines and tissues. Taking Enformer as the feature extractor, for each input DNA sequence of 196,608 bp, the Enformer model generates an *d* = 5,313-dimensional feature vector for each of the central non-overlapping *k* = 896 bins, each with a bin size of 128 bp. B) Sliding Window Approach: Given that a single linkage disequilibrium (LD) block is typically larger than the input length for Enformer, we utilize a sliding window approach within each LD block to capture features from multiple input regions. If an LD block contains *l* input regions, the extracted features for this block will have dimensions of (*l, k, d*). We denote these features as 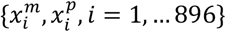, where the superscript indicates the parent of origin (*m* for maternal and *p* for paternal). C) Dimension Reduction: To manage the high dimensionality of the extracted features, we apply local Principal Component Analysis (PCA) to each bin across individuals. This dimension reduction step reduces the features to a more manageable size, denoted as *d*^′^ where *d*^′^≪ *d*.

#### Risk Prediction

The final step involves building predictive models for disease risk based on the reduced-dimension features and the phenotypic data: A) Model Construction: For binary classification tasks, such as disease presence or absence, we construct a GBRT classification model. For continuous traits, we use GBRT regression model. These models are trained on the individual features obtained from the feature extraction step. B) Training and Testing: We randomly select 80% of both case and control subjects as the training set, leaving the remaining 20% as the testing set. The performance of the models is evaluated using prediction results on the testing set. C) Performance Evaluation: The predictive accuracy of the models is assessed using metrics such as the AUC for classification tasks and the coefficient of determination (R^2^) for regression tasks. These metrics provide insight into the effectiveness of Epi-PRS in predicting disease risk based on integrated genomic and epigenomic data.

### Genomic LLM

Different from previous convolutional and recurrent neural network models, recent developments in LLMs typically rely on the transformer architecture, which was introduced by Vaswani *et al*. (46) as a revolutionized deep learning method, offering more efficient training and better handling of long-range dependencies in sequential data. The vanilla transformer model is divided into two main components: the encoder and the decoder, both of which share a similar basic architecture composed of a stack of identical blocks (47). Each block in the transformer model consists of two key sub-layers: 1) Multi-Head Attention Sub-Layer: This sub-layer allows the model to attend to different positions of the input sequence simultaneously, capturing various aspects of the data. It computes attention scores in multiple parallel heads, providing diverse perspectives on the input data. These scores are then aggregated to form a comprehensive understanding of the sequence.

Feed-Forward Sub-Layer: This sub-layer consists of fully connected feed-forward networks applied independently to each position in the sequence. It includes a non-linear activation function, typically ReLU, which helps in capturing complex patterns and interactions within the data. Both sub-layers are followed by layer normalization, which standardizes the inputs to each layer, improving the stability and performance of the model. Additionally, a residual connection (48) around every sub-layer is applied in each block to help mitigate the vanishing gradient problem. These residual connections add the input of the sub-layer to its output, ensuring that the gradients can flow through the network more effectively during backpropagation.

Our previous review (49) describes each module and layer that constitutes the transformer model in detail, exploring their mechanisms and potential applications in bioinformatics. In this paper, we adopt Enformer (50), one of the most advanced genomic LLM models, as our default choice of LLM. Enformer innovatively integrates the transformer encoder structure to predict 5,313 epigenomic signals in humans, significantly enhancing its capabilities over previous CNN-based models. Enformer brings several improvements that make it particularly well-suited for genomic data: 1) Extended Receptive Field: The transformer encoder structure in Enformer greatly increases the receptive field of the network to 196,608 bp. This extensive receptive field allows the model to capture long-range interactions and dependencies within the genomic sequence, which are crucial for understanding complex regulatory mechanisms and gene expression patterns. 2) Improved Predictive Performance: By leveraging the transformer architecture, Enformer achieves superior predictive performance in estimating a wide array of epigenomic signals, including chromatin accessibility, transcription factor binding, and histone modifications. These improvements make Enformer highly effective in modeling the intricate relationships between genetic variants and molecular phenotypes.

Overall, the adoption of Enformer in our study exemplifies the significant advancements in genomic LLMs, enabling more accurate and comprehensive predictions of epigenomic features from genomic sequences. This innovation is a key component of the Epi-PRS framework, which leverages these detailed molecular phenotype predictions to enhance disease risk prediction and improve our understanding of the genetic architecture of complex traits.

### Dimension reduction of genomic LLM features

For each 196,608 bp input region, the Enformer-based method generates a substantial number of predicted features—approximately 9.5 million (896 bins × 5,313 features per bin × 2 haploid genomes). This high dimensionality presents a significant challenge for risk modeling, as traditional statistical methods struggle to handle such a vast number of predictor variables effectively. To address this issue, we implement local PCA strategies for dimension reduction, which is demonstrated in the four steps below.

#### 1. Feature Vector Formation

For the *i*^*th*^ bin, we construct a set of feature vectors 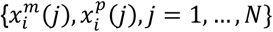 where *j* indexes all cases or controls in the study, and 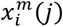 and 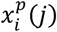 represent the maternal and paternal feature vectors, respectively. Each feature vector contains 5,313 dimensions, corresponding to the genomic and epigenomic signals predicted by Enformer.

#### 2. Principal Component Analysis (PCA)

PCA is performed on this set of feature vectors to reduce the dimensionality from 5,313 to a much smaller value. This reduction is crucial for making the subsequent risk modeling computationally feasible and statistically robust. For example, we reduce the dimension to 5 principal components, which capture the majority of the variance in the original high-dimensional feature vectors.

#### 3. Reduced-Dimension Feature Vectors

After PCA, we obtain a sequence of reduced-dimension feature vectors 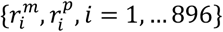 for each individual in the study. These vectors characterize the epigenomic states of the input region while retaining the most informative aspects of the original data. Here, 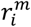 and 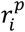 are the reduced-dimension feature vectors for the maternal and paternal sequences, respectively, for the *i*^*th*^bin.

#### 4. Combining Features Across Bins

The reduced-dimension feature vectors from all 896 bins are then concatenated to form a comprehensive feature set for each individual. This results in a significantly reduced yet highly informative feature matrix, which captures the essential epigenomic signals across the entire 196,608 bp input region.

By adopting local PCA for dimension reduction, we can manage the high dimensionality of the Enformer-predicted features, ensuring that our risk models are both computationally efficient and statistically robust. This approach allows us to harness the rich epigenomic information provided by Enformer, ultimately leading to more accurate and comprehensive disease risk predictions.

### Selection of informative LD block and application in UKBB

In principle, we could include the reduced-dimension features from each approximately 200 kb region tiling the genome as predictor variables in the risk model. However, given that there are about 15,000 such regions across the genome, each contributing 4,480 features (896 bins × 5 principal components), the number of predictor variables could still be prohibitively large. To address this issue, we propose a further dimension reduction strategy by focusing on regions pre-selected based on the enrichment of GWAS signals for the phenotype of interest. We regard the presence of SNPs with highly significant *p*-values for association with the disease as an indication that the corresponding LD block contains genetic variations useful for risk modeling.

We can summarize the main steps of our method as follows:

1. **Joint Variant Calling and Phasing**:
  a. Perform joint variant calling for the individuals in the study based on their WGS data.
  b. Phase the detected SNVs to obtain haploid genome sequences for each individual, distinguishing maternal and paternal genomes.
2. **GWAS-Based SNP Selection**:
  a. Utilize prior GWAS studies or perform GWAS analysis using the current dataset to identify a set of SNPs associated with the disease of interest. These SNPs should have *p*-values smaller than a predefined threshold (denoted as *p*_0_).
  b. Select all LD blocks that contain SNPs with p-values less than *p*_0_. This ensures that we focus on genomic regions that are most likely to contribute to the disease risk.
3. **Tiling LD Blocks and Feature Extraction**:
  a. For each selected LD block, tile the block with overlapping 196,608 bp input regions, focusing on the central 114,668 bp region to ensure comprehensive coverage.
  b. Apply the Enformer model to the maternal and paternal input sequences for these regions to obtain the epigenomic feature vectors 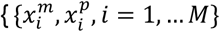, where *M* indicates the total number of 128 bp bins required to cover the LD block.
4. **Dimension Reduction of Epigenomic Features**:
  a. Perform PCA on the feature vectors 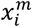, 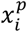 for each bin to reduce the dimensionality from 5,313 to a smaller value (e.g., 5 principal components).
  b. Concatenate the reduced-dimension feature vectors 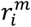,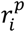 across all bins within each LD block to form a comprehensive feature vector for the entire LD block.
5. **Risk Prediction Modeling**:
  a. Construct a feature vector *x*_*b*_ for each selected LD block.
  b. Concatenate the PCA-projected feature vectors 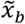 from all selected LD blocks to form a global feature vector 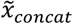.
  c. Apply GBRT or other binary prediction methods to learn a risk predictor based on 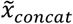. Alternatively, fit LD block-specific risk prediction models based on 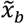 for each LD block and then learn an optimally weighted combination of these models as the final risk prediction model.
6. **Application to UK Biobank Data**:
  a. Obtain GWAS summary statistics data from the UKBB, excluding the testing individuals for each disease, to identify the most significant variants for breast cancer and diabetes.
  b. Apply a stringent p-value cutoff of 5*e* − 17 for breast cancer, identifying 89 significant variants, and a cutoff of 5*e* − 20 for diabetes, identifying 3,015 significant variants.
  c. Intersect these significant variants with 1,702 LD blocks and identify 5 LD blocks for breast cancer and 11 LD blocks for diabetes that contain significant variants.

By integrating GWAS-enriched regions and performing local PCA for dimension reduction, our approach effectively narrows down the predictor variables to the most relevant genomic features. This enables more efficient and accurate risk modeling, leveraging both genotypic and epigenomic information to provide a comprehensive understanding of disease susceptibility.

### Phenotype simulation

To systematically evaluate the performance, we aim to simulate phenotypes using real genotype data from the UK BioBank. Since Epi-PRS integrates regulatory information, which is cell-type specific, we develop a regulatory-based phenotype simulation tool that can sample both rare and common variants. This allows us to control the fraction of rare variants contributing to the disease phenotype, providing a comprehensive evaluation of Epi-PRS across different genetic architectures.

For *n* individuals, let *y*_*i*_ denote a binary trait (1 indicating a case and 0 indicating a control) following the Bernoulli distribution with mean *μ*_*i*_, *i* = 1, …, *n*. We consider the following generalized linear model:

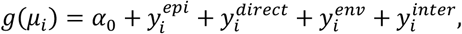

where *g* is the logit function,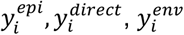 and 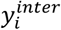 denote the epigenetic effect, SNP direct effect, environmental effect and SNP interaction effect, respectively. *α*_0_ is determined such that the expected number of cases equals *n*λ where λ is the pre-specified sample prevalence rate and is set to 0.5.

#### Epigenetic Effect

We assume 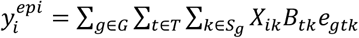. Here, *G* is the set of causal genes and we randomly selected one causal gene per LD block. *T* is the set of expressed transcription factors (TFs) with median reads > 10 in blood tissue from GTEx. *S*_*g*_ is the set of causal variants in neighboring regulatory elements (REs) for gene *g* (distance to the gene body < 1MB). REs are defined as candidate cis-regulatory elements (cCREs) in blood tissue from ENCODE, and we consider 100 causal variants per causal gene throughout the simulations. *X*_*i*1_ denotes the normalized genotype for variant *k* in individual *i. B*_*tk*_ is a binary variable representing whether variant *k* overlaps with binding site of TF *t* where 1 indicates overlap and 0 indicates no overlap. TF binding sites are derived using the motif scanning tool HOMER to scan along the reference genome and match the motifs of each TF. We further assume the random effect term 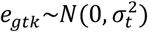, where 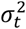 follows an inverse gamma distribution *IG*(3, 1). The epigenetic effect 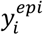 is then normalized to have mean 0 and variance 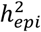 across *n* individuals. Cases and controls are simulated by thresholding in these *n* individuals.

#### SNP Direct Effect

We assume 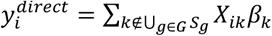, where β_1_∼*N*(0,1). The SNP direct effect is summed over SNPs that do not overlap any REs of causal genes. Here, we assume that the epigenetic effect is independent of the SNP direct effect, which is convenient for evaluation. The SNP direct effect is further normalized to have mean 0 and variance *h*^7^ across *n* individuals.

#### Environment Effect

We assume 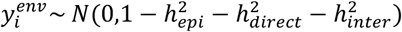

#### SNP Interactions Effect

We assume a fraction of the causal SNPs have multiplicative interaction effect (where one SNP increases or decreases the effect of the other).

By combining these effects, we can simulate phenotypes that reflect the complex interplay among three components: 1) genetic variants, 2) regulatory elements, 3) environmental factors and 4) SNP interactions. Additionally, we can adjust the proportion of rare variants contributing to the disease by selectively sampling these variants during the simulation process. This flexible simulation approach allows us to comprehensively assess the robustness and accuracy of Epi-PRS in various genetic contexts, providing valuable insights into its performance across different scenarios. Note that the Epi-PRS method implicitly assumes a model in which genetic variants affect the phenotype through their effects on epigenomic features. Instead of simulating phenotype based on such a model, we opt to use the above random effect model in order to avoid biasing the comparison in favor of our method. For each setting considered, the simulation was repeated 20 times.

## Data Availability

All data produced in the present study are available upon reasonable request to the authors

## Data availability

Raw data (genotype and phenotype) from the UKBB participants can be requested from the UKBB Access Management System (https://bbams.ndph.ox.ac.uk).

## Code availability

Code is publicly available online at https://github.com/SUwonglab/Epi-PRS.

## Acknowledgments

The works of Z.W., H.G., Q.L., and W.H.W. were partially supported by NIH grants R01 HG010359, P50 HG007735 and HG00773506. The work of Q.L. was also supported by a NIH grant K99HG013661.

