## Supplementary for "How to improve polygenic prediction from whole-genome sequencing data by leveraging predicted epigenomic features?"

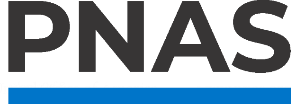

**Supporting Information for**

How to improve polygenic prediction from whole-genome sequencing data by leveraging predicted epigenomic features?

Wanwen Zeng^1,4^, Hanmin Guo^1,3,4^, Qiao Liu^1,4^, and Wing Hung Wong^1,2,4,*^

^1^ Department of Statistics, Stanford University, Stanford, CA 94305, USA;

^2^ Department of Biomedical Data Science, Stanford University, Stanford, CA 94305, USA;

^3^ Department of Psychiatry and Behavioral Sciences, Stanford University, Stanford, CA 94305, USA;

^4^ Bio-X Program, Stanford University, Stanford, CA 94305, USA;

* To whom correspondence should be addressed

**This PDF file includes:**

Tables S1 to S6

**Table S1. Simulated Results with Varying the Number of Training Data**. This table presents the simulated results for the genomic region chr6:192074807-21684054, highlighting the impact of linear and non-linear models with varying training data set. The evaluation metric used is the average AUC score, calculated over 20 repetitions. In these simulations, the proportion of epigenetic effects and the propitiation of rare variants are set to zero.

| chr6:192074807-21684054 | 1,000 | 2,000 | 4,000 | 8,000 | 16,000 |
| --- | --- | --- | --- | --- | --- |
| Epi-PRS | 0.5886 ± 0.0167 | **0.6256 ± 0.0190** | **0.6605 ± 0.0196** | **0.6866 ± 0.0112** | 0.7126 ± 0.0115 |
| PRS-CS (without additional GWAS) | **0.6127 ± 0.0301** | 0.6160 ± 0.0157 | 0.6000 ± 0.0273 | 0.6421 ± 0.0329 | 0.6548 ± 0.0175 |
| LDPred2 (without additional GWAS) | 0.5912 ± 0.0411 | 0.6230 ± 0.0387 | 0.5999 ± 0.0356 | 0.6511 ± 0.0397 | 0.6692 ± 0.0465 |
| Genotype-PCA-GBRT | 0.5593 ± 0.0242 | 0.5971 ± 0.0255 | 0.6171 ± 0.0448 | 0.6707 ± 0.0308 | 0.7213 ± 0.0259 |
| Genotype-GBRT | 0.5828 ± 0.0267 | 0.6139 ± 0.0202 | 0.6595 ± 0.0268 | 0.6578 ± 0.0193 | **0.7324 ± 0.0291** |

**Table S2. Simulated Results with Varying the Number of Training Data**. This table presents the simulated results for the genomic region chr6:31571218-32682664, highlighting the impact of linear and non-linear models with varying training data set. The evaluation metric used is the average AUC score, calculated over 20 repetitions. In these simulations, the proportion of epigenetic effects and the propitiation of rare variants are set to zero.

| chr6:31571218-32682664 | 1,000 | 2,000 | 4,000 | 8,000 | 16,000 |
| --- | --- | --- | --- | --- | --- |
| Epi-PRS | 0.5915 ± 0.0169 | 0.6390 ± 0.0184 | **0.6775 ± 0.0213** | **0.7062 ± 0.0191** | 0.7360 ± 0.0172 |
| PRS-CS (without additional GWAS) | 0.5919 ± 0.0220 | 0.6220 ± 0.0121 | 0.6316 ± 0.0189 | 0.6528 ± 0.0209 | 0.6899 ± 0.0142 |
| LDPred2 (without additional GWAS) | **0.5975 ± 0.0204** | 0.6208 ± 0.0265 | 0.6349 ± 0.0227 | 0.6625 ± 0.0172 | 0.6761 ± 0.0153 |
| Genotype-PCA-GBRT | 0.5860 ± 0.0168 | 0.6426 ± 0.0252 | 0.6664 ± 0.0126 | 0.7047 ± 0.0171 | 0.7249 ± 0.0193 |
| Genotype-GBRT | 0.5815 ± 0.0185 | **0.6448 ± 0.0184** | 0.6679 ± 0.0175 | 0.7015 ± 0.0208 | **0.7392 ± 0.0209** |

**Table S3.** **Simulated Results with Varying Proportions of Epigenetic Effects**. This table presents the simulated results for the genomic region chr6:192074807-21684054, highlighting the impact of different proportions of epigenetic effects on prediction performance. The evaluation metric used is the average AUC score, calculated over 20 repetitions. In these simulations, the proportion of rare variants is set to zero.

| chr6:192074807-21684054 | 0% | 25% | 50% | 75% | 100% |
| --- | --- | --- | --- | --- | --- |
| Epi-PRS | **0.7115 ± 0.0176** | 0.7160 ± 0.0184 | **0.7290 ± 0.0160** | 0.7555 ± 0.0193 | **0.7783 ± 0.0217** |
| Epi-PRS (blood features only) | 0.6861 ± 0.0191 | 0.7070 ± 0.0199 | 0.7292 ± 0.0196 | **0.7735 ± 0.0224** | 0.7691 ± 0.0196 |
| PRS-CS (without additional GWAS) | 0.6649 ± 0.0211 | 0.6707 ± 0.0223 | 0.6625 ± 0.0235 | 0.6716 ± 0.0217 | 0.6724 ± 0.0199 |
| LDPred2 (without additional GWAS) | 0.6603 ± 0.0174 | 0.6801 ± 0.0202 | 0.6816 ± 0.0184 | 0.6794 ± 0.0180 | 0.6709 ± 0.0196 |
| Genotype-PCA-GBRT | 0.7044 ± 0.0201 | **0.7169 ± 0.0203** | 0.7170 ± 0.0159 | 0.7150 ± 0.0231 | 0.7178 ± 0.0129 |
| Genotype-GBRT | 0.7035 ± 0.0231 | 0.7109 ± 0.0160 | 0.7238 ± 0.0189 | 0.7066 ± 0.0135 | 0.7157 ± 0.0219 |

**Table S4.** **Simulated Results with Varying Proportions of Epigenetic Effects**. This table presents the simulated results for the genomic region chr6:31571218-32682664, highlighting the impact of different proportions of epigenetic effects on prediction performance. The evaluation metric used is the average AUC score, calculated over 20 repetitions. In these simulations, the proportion of rare variants is set to zero.

| chr6:31571218-32682664 | 0% | 25% | 50% | 75% | 100% |
| --- | --- | --- | --- | --- | --- |
| Epi-PRS | 0.7357 ± 0.0180 | **0.7553 ± 0.0216** | **0.7656 ± 0.0149** | **0.7785 ± 0.0183** | 0.7817 ± 0.0183 |
| Epi-PRS (blood features only) | 0.7344 ± 0.0182 | 0.7432 ± 0.0192 | 0.7619 ± 0.0174 | 0.7776 ± 0.0246 | **0.7834 ± 0.0208** |
| PRS-CS (without additional GWAS) | 0.6889 ± 0.0180 | 0.6842 ± 0.0167 | 0.6796 ± 0.0149 | 0.6779 ± 0.0170 | 0.6782 ± 0.0159 |
| LDPred2 (without additional GWAS) | 0.6736 ± 0.0192 | 0.6813 ± 0.0142 | 0.6813 ± 0.0169 | 0.6831 ± 0.0184 | 0.6804 ± 0.0245 |
| Genotype-PCA-GBRT | 0.7234 ± 0.0205 | 0.7386 ± 0.0202 | 0.7325 ± 0.0187 | 0.7494 ± 0.0220 | 0.7403 ± 0.0195 |
| Genotype-GBRT | **0.7457 ± 0.0187** | 0.7413 ± 0.0178 | 0.7330 ± 0.0198 | 0.7358 ± 0.0224 | 0.7447 ± 0.0240 |

**Table S5.** **Simulated Results with Varying Proportions of Rare Variants.** This table presents the simulated results for the genomic region chr6:192074807-21684054, highlighting the impact of different proportions of rare variants on prediction performance. When the proportion of rare variants increases to 100%, traditional PRS methods fail to predict effectively as they only consider common variants. The evaluation metric used is the average AUC score, calculated over 20 repetitions. In these simulations, the proportion of epigenetic effect is set to zero.

| chr6:192074807-21684054 | 0% | 25% | 50% | 75% | 100% |
| --- | --- | --- | --- | --- | --- |
| Epi-PRS | **0.7486 ± 0.0154** | **0.7477 ± 0.0192** | **0.7522 ± 0.0192** | 0.7362 ± 0.0166 | **0.7510 ± 0.0246** |
| PRS-CS (without additional GWAS) | 0.6169 ± 0.0183 | 0.6208 ± 0.0219 | 0.5668 ± 0.0195 | 0.5253 ± 0.0210 | NA |
| LDPred2 (without additional GWAS) | 0.6137 ± 0.0166 | 0.5945 ± 0.0189 | 0.5506 ± 0.0177 | 0.5215 ± 0.0232 | NA |
| Genotype-PCA-GBRT | 0.7410 ± 0.0191 | 0.7454 ± 0.0198 | 0.7433 ± 0.0263 | **0.7406 ± 0.0179** | 0.7450 ± 0.0170 |
| Genotype-GBRT | 0.7422 ± 0.0193 | 0.7423 ± 0.0177 | 0.7382 ± 0.0180 | 0.7368 ± 0.0222 | 0.7444 ± 0.0184 |

**Table S6.** **Simulated Results with Varying Proportions of Rare Variants.** This table presents the simulated results for the genomic region chr6:31571218-32682664, highlighting the impact of different proportions of rare variants on prediction performance. When the proportion of rare variants increases to 100%, traditional PRS methods fail to predict effectively as they only consider common variants. The evaluation metric used is the average AUC score, calculated over 20 repetitions. In these simulations, the proportion of epigenetic effect is set to zero.

| chr6:31571218-32682664 | 0% | 25% | 50% | 75% | 100% |
| --- | --- | --- | --- | --- | --- |
| Epi-PRS | 0.7420 ± 0.0196 | **0.7650 ± 0.0152** | **0.7506 ± 0.0171** | 0.7518 ± 0.0218 | **0.7594 ± 0.0171** |
| PRS-CS (without additional GWAS) | 0.6244 ± 0.0174 | 0.6196 ± 0.0233 | 0.5697 ± 0.0157 | 0.5557 ± 0.0135 | NA |
| LDPred2 (without additional GWAS) | 0.6386 ± 0.0247 | 0.5993 ± 0.0205 | 0.5598 ± 0.0221 | 0.5467 ± 0.0148 | NA |
| Genotype-PCA-GBRT | 0.7263 ± 0.0209 | 0.7538 ± 0.0171 | 0.7385 ± 0.0177 | 0.7501 ± 0.0199 | 0.7505 ± 0.0209 |
| Genotype-GBRT | **0.7556 ± 0.0285** | 0.7646 ± 0.0232 | 0.7351 ± 0.0176 | **0.7596 ± 0.0203** | 0.7516 ± 0.0181 |
